# Subregional analysis of the amygdala, thalamus, and hypothalamus at the pre-decline stage in Parkinson’s disease groups with later cognitive impairment

**DOI:** 10.1101/2024.12.18.24319260

**Authors:** Kazuhide Seo, Genko Oyama, Toshimasa Yamamoto

## Abstract

While cognitive decline in Parkinson’s disease (PD) significantly impacts patients’ quality of life, early detection remains challenging. Recent advances in magnetic resonance imaging analysis have enabled detailed examination of subcortical structures. This study aimed to investigate subtle changes in specific subregions of the amygdala, thalamus, and hypothalamus in PD patients before they developed cognitive decline. Magnetic resonance imaging data of 163 participants (97 healthy controls [HC], 45 PD patients with normal cognition [PDNC], and 21 PD patients who show cognitive impairment [PDCI]) from the Parkinson’s Progression Markers Initiative database were analyzed. Detailed subregional analyses of brain structures were performed. Cognitive function was assessed using the Montreal Cognitive Assessment and domain-specific tests. The PDCI group exhibited significantly lower intracranial occupancy rates in specific subregions of the amygdala, thalamus, and hypothalamus than the HC group; however, these changes did not correlate significantly with cognitive test scores. Conversely, significant structural changes were observed in extensive cortical regions, subcortical gray matter areas, and white matter areas, which correlated with various cognitive functions including memory, attention, executive function, and visuospatial abilities. Nevertheless, no significant associations were found between changes in individual brain regions and the risk of mild cognitive impairment progression. This study elucidates early brain structural changes associated with cognitive decline in PD. While structural alterations were observed in the amygdala, thalamus, and hypothalamus, widespread cortical changes demonstrated stronger associations with cognitive decline. These findings suggest that cognitive impairment in PD results from extensive cortical network alterations rather than changes in specific subcortical regions. This insight emphasizes the need for a comprehensive approach, considering multiple brain regions and their interactions, in early diagnosis and intervention strategies for PD-related cognitive impairment.

## Introduction

Parkinson’s disease (PD) is a progressive neurodegenerative disorder characterized by both motor and non-motor symptoms [1]. Among the non-motor symptoms, cognitive impairment is particularly common and significantly impacts patients’ quality of life [2]. The average prevalence of mild cognitive impairment (MCI) in PD is 27% (range, 19%–38%), and it is associated with an increased risk of developing dementia in PD [3]. Cognitive decline in PD is considered multifactorial, involving various neurotransmitter systems and pathological processes, including dopaminergic, cholinergic, and noradrenergic deficits [4], as well as the accumulation of alpha-synuclein and beta-amyloid proteins [5].

The Movement Disorder Society (MDS) has established diagnostic criteria for PD-MCI [3]. These criteria include two levels of assessment: Level I criteria, which allow for a less comprehensive cognitive evaluation using global cognitive scales or a limited battery of neuropsychological tests, and Level II criteria, which require more detailed neuropsychological testing covering five cognitive domains. While both levels are validated diagnostic tools, they primarily rely on neuropsychological tests and may not capture the earliest stages of cognitive decline [3]. Although Level II criteria provide a more comprehensive assessment, the extensive testing required can be impractical in many clinical settings. Furthermore, there is a need for more convenient and sensitive diagnostic tools that can detect cognitive impairment before it becomes clinically apparent because early detection is crucial in planning appropriate care strategies and aids and in evaluating candidacy of invasive treatments such as deep brain stimulation.

Quantitative analysis techniques of brain MRI have made it possible to capture subtle changes in brain structure. In particular, methods such as voxel-based morphometry, cortical thickness analysis, and diffusion tensor imaging (DTI) allow for quantitative assessment of gray matter volume, cortical thickness, and white matter microstructure [6]. These techniques can detect early brain structural changes that are difficult to identify through conventional visual assessment, potentially aiding in the early detection and prediction of cognitive impairment progression [7].

Regarding brain structural changes associated with cognitive decline in PD, multiple meta-analyses have revealed patterns of gray matter atrophy in PD-MCI, primarily affecting the frontal, temporal, and parietal lobes, as well as the insula and limbic system [8–11]. Furthermore, studies focusing on PD-MCI converters have shown cortical thinning at baseline in the anterior cingulate, parietal, temporal, and occipital cortices. These studies have also documented atrophy in the nucleus accumbens and progressive volume loss in the thalamus, caudate nucleus, nucleus accumbens, and hippocampal CA2-3 regions [12–16]. These findings suggest that brain structural changes beginning in the cognitively normal stage are associated with subsequent transition to MCI.

Recent advances in brain MRI analysis techniques have enabled the analysis of fine subregions in areas such as the hippocampus [17], amygdala [18], thalamus [19], and hypothalamus [20]. This provides an opportunity to capture subtle structural changes in these regions and better understand the mechanisms of cognitive decline in PD. Several studies have already conducted detailed subregional analyses of the hippocampus. For instance, PD-MCI patients have shown volume reductions in CA1 and the hippocampal–amygdaloid transition area (HATA) compared with healthy controls (HCs) and PD–normal cognitive impairment (NCI) patients [21]. Moreover, the volumes of hippocampal subregions, particularly the hippocampal para-gyrus, CA4, granule cell layer, and HATA regions, have been shown to be important predictors of transition from PD-NCI to PD-MCI [16, 22].

However, few studies have conducted detailed analyses of the subregions of the amygdala, thalamus, and hypothalamus to investigate structural abnormalities prior to MCI transition. The amygdala shows accumulation of Lewy bodies from early stages, leading to dysfunction in emotional and memory processing [23, 24]. Impairment of cholinergic neurotransmission in the thalamus is thought to play a crucial role in the onset and progression of cognitive symptoms in PD [25]. The hypothalamus in PD exhibits widespread Lewy body pathology [26], resulting in reduced functional connectivity with other brain regions. This pathological change leads to dysregulation of the autonomic nervous system, affecting blood pressure control and thermoregulation [27]. Such autonomic dysfunction may indirectly impact cognitive function, possibly through changes in cerebral hemodynamics [28]. Thus, the predictive value of subregions within the amygdala, thalamus, and hypothalamus for PD-MCI has not yet been elucidated.

The main focus of this study is to leverage recent advances in MRI analysis techniques to examine fine subregions of the amygdala, thalamus, and hypothalamus—structures that have been technically challenging to analyze in detail until now. We aim to investigate whether structural abnormalities in these specific subregions exist before PD transitions to MCI. This comprehensive approach may enable us to capture more precise patterns of brain structural changes from the early stages of cognitive decline in PD, potentially contributing to early diagnosis and prediction of progression.

## Materials and Methods

### Research Participants

All MRI and clinical data used in this study were obtained from the Parkinson’s Progression Markers Initiative (PPMI) database (https://www.ppmi-info.org/). According to the PPMI cohort criteria, PD patients at baseline had to meet the following conditions: (i) have at least two of the following: resting tremor, bradykinesia, rigidity, or either asymmetric resting tremor or asymmetric bradykinesia; (ii) be in an early clinical stage (within 2 years of PD diagnosis and have Hoehn and Yahr [H&Y] stage I or II); (iii) not be taking PD medication; (iv) have confirmed dopamine transporter (DAT) deficiency on imaging; (v) be judged as not having dementia by the research site investigator; (vi) have a 15-item Geriatric Depression Scale (GDS-15) score < 5. The HC group met the following criteria: (i) no significant neurological dysfunction and (ii) no first-degree relatives with PD.

From the PPMI cohort, we selected participants meeting our MRI criteria and further excluded those with (i) poor image quality, (ii) history of cerebrovascular disease or head trauma that could affect brain image analysis, (iii) baseline Montreal Cognitive Assessment (MoCA) score < 26, or (iv) missing baseline clinical data. The final sample consisted of 163 participants: 97 HC and 66 PD patients.

PD patients were further divided into two groups based on their cognitive status during the 4-year follow-up period: 21 PD patients who show cognitive impairment (PDCI) and 45 PD patients who maintained normal cognitive function (PDNC). For cognitive assessment, we utilized MoCA, which has been widely established as a validated assessment tool for cognitive function in Parkinson’s disease even before the publication of the MDS Task Force guidelines [29]. While Level II diagnosis in the guidelines enables comprehensive evaluation using multiple tests across five cognitive domains (attention/working memory, memory, language, executive function, and visuospatial function), such extensive neuropsychological testing is often impractical in clinical settings owing to time constraints. Studies have demonstrated that Level I criteria, including MoCA, show comparable effectiveness to Level II criteria in predicting transition to PD-MCI [30], and MoCA specifically has proven reliable in predicting long-term cognitive outcomes [31]. Therefore, PDCI was defined as participants whose MoCA score fell below 26 during the follow-up period, following Level I of the MDS PD-MCI diagnostic criteria [3]. This approach aligns with current clinical practice and has been widely adopted in predictive modeling studies of cognitive decline in PD [32]. Table 1 shows the number of PDCI during each year of the follow-up period. The cumulative number of PDCI cases over the 4-year follow-up is presented.

**Table 1.** Incidence of PDCI over the 4-year follow-up period. This table illustrates the number of Parkinson’s disease patients who show cognitive impairment (PDCI) during each year of the 4-year follow-up period. In the first year, the number of PDCI is 12, followed by 6 in the second year and 3 in the third year. No PDCI are present in the fourth year. The total number of PDCI over the entire follow-up period is 21.

|  | 1 year | 2 year | 3 year | 4 year | Total |
| --- | --- | --- | --- | --- | --- |
| **PDCI** | 12 | 6 | 3 | 0 | 21 |

### Study Design

This study adopted a longitudinal design. All participants underwent baseline MRI scans and clinical evaluations. PD patients received annual follow-up examinations for 4 years, with repeated cognitive function assessments using MoCA.

### MRI

All MRI examinations were conducted using a 3.0T MRI scanner (Siemens Healthcare, USA) following a standard protocol. A 3D magnetization-prepared rapid gradient-echo sequence was used for brain anatomy imaging (176 axial slices, repetition time = 2300 ms, echo time = 2.98 ms, flip angle = 9°, voxel size 1 mm × 1 mm × 1 mm). A gradient-echo echo-planar imaging sequence over 210 volumes or time points was used for resting-state brain functional activity imaging (40 axial slices, repetition time = 2400 ms, echo time = 25 ms, flip angle = 80°, voxel size 3.3 mm × 3.3 mm × 3.3 mm). Detailed information can be found at https://www.ppmi-info.org/study-design/researchdocuments-and-sops/.

### Image Analysis

T1-weighted images were processed using FreeSurfer (version 7.3.2, https://surfer.nmr.mgh.harvard.edu/) for automatic segmentation and volumetry of brain regions. Subregional analyses of the hippocampus, amygdala, thalamus, subthalamic nucleus, and brainstem were performed. The brain was segmented into 68 cortical regions, 16 deep gray matter regions, 70 white matter regions, 4 cerebellar regions, 38 hippocampal regions, 18 amygdala regions, 50 thalamic regions, 10 hypothalamic regions, and 4 brainstem regions.

The hippocampus, amygdala, thalamus, and hypothalamus were divided into the following regions for both left and right hemispheres:

Hippocampus (19 regions): Cornu Ammonis 1-body (CA1-body), CA1-head, CA3-body, CA3-head, CA4-body, CA4-head, fimbria, granule cells-molecular layer-dentate gyrus-body (GC-ML-DG-body), GC-ML-DG-head, HATA, hippocampal tail, hippocampal fissure, molecular layer-hippocampus-body (molecular_layer_HP-body), molecular_layer_HP-head, parasubiculum, presubiculum-body, presubiculum-head, subiculum-body, subiculum-head.

Amygdala (9 regions): accessory basal nucleus, anterior amygdaloid area, basal nucleus, central nucleus, cortical nucleus, corticoamygdaloid transition area, lateral nucleus, medial nucleus, paralaminar nucleus.

Thalamus (25 regions): anteroventral nucleus, central medial nucleus, central lateral nucleus, centromedian nucleus, lateral segment of the suprageniculate nucleus, lateral dorsal nucleus, lateral geniculate nucleus (LGN), lateral posterior nucleus, medial dorsal nucleus lateral part, medial dorsal nucleus medial part (MDm), medial geniculate nucleus, medial ventral nucleus (reuniens), paracentral nucleus, parafascicular nucleus, paratenial nucleus, pulvinar anterior part (PuA), pulvinar inferior part, pulvinar lateral part, pulvinar medial part, ventral anterior nucleus, ventral anterior nucleus magnocellular part, ventral lateral nucleus anterior part, ventral lateral nucleus posterior part, ventral medial nucleus, ventral posterolateral nucleus.

Hypothalamus (5 regions): anterior-inferior, anterior-superior, posterior, inferior tubular, superior tubular.

This detailed segmentation allowed for the assessment of subtle structural changes in each brain region. Clinical and neuropsychological assessments disease stage was evaluated using the H&Y scale, and motor symptoms were assessed using the MDS–Unified Parkinson’s Disease Rating Scale (MDS-UPDRS). Depressive symptoms were evaluated using GDS-15. Cognitive function was assessed using MoCA and domain-specific tests. Domain-specific tests included the Hopkins Verbal Learning Test-Revised for memory, Benton Judgment of Line Orientation 15-item (half) version for visuospatial function, Symbol-Digit Modalities Test (SDMT) for processing speed and attention, and Letter-Number Sequencing (LNS) and semantic fluency (animals) for executive function and working memory.

### Statistical Analysis

Statistical analyses were performed using JMP pro version 16.2.0 (SAS Institute Inc., Cary, NC, USA). Demographic and clinical information were analyzed using analysis of covariance for quantitative data and Pearson’s chi-square test for qualitative data.

### Comparison of Intracranial Occupancy Rates

In brain volume analysis, intracranial occupancy rates were calculated by dividing measured volumes by intracranial volume. Comparisons of intracranial occupancy rates between groups were performed using analysis of covariance adjusted for age, sex, handedness, and education level.

### Association Between Cognitive Test Scores and Brain Regions

Analysis of the association between brain region volumes and cognitive test scores was conducted for brain regions showing significant differences in intracranial occupancy rates between PDCI and PDNC groups. This analysis used analysis of covariance adjusted for age, sex, handedness, and education level.

### Hazard Ratios for MCI Progression

For regions showing significant differences between PDCI and PDNC, Z-scores of intracranial occupancy rates were calculated using the entire PD sample as the reference population. Survival analysis using Cox proportional hazards model was performed to calculate hazard ratios for MCI progression based on these Z-scores. The event was defined as the point when the MoCA score fell below 26. The model was adjusted for age, sex, handedness, and education level.

### Ethical Considerations

All PPMI sites obtained approval from their ethics standards committees before the study began, and all participants provided written informed consent before participation. Detailed information on PPMI protocol approval, registration, and patient consent can be found at https://www.ppmi-info.org/.

## Results

### Demographic and Clinical Characteristics

As shown in Table 2, the PDCI group had a significantly higher proportion of men than the HC group (p = 0.0362) and PDNC group (p = 0.0166) and was significantly older than the PDNC group (62.6 ± 8.0 vs 57.8 ± 9.8 years, p = 0.0463). Regarding cognitive function, the PDCI group showed significantly lower scores than the HC and PDNC groups in HVLT-R immediate recall (PDCI: 22.9 ± 4.0, HC: 25.9 ± 4.4, PDNC: 27.8 ± 4.6; p < 0.01), delayed recall (PDCI: 7.5 ± 2.7, HC: 9.2 ± 2.2, PDNC: 10.0 ± 1.6; p < 0.01), LNS (PDCI: 10.0 ± 2.2, HC: 11.4 ± 2.6, PDNC: 11.7 ± 1.6; p < 0.05), and SDMTOTAL (PDCI: 37.4 ± 8.4, HC: 48.0 ± 10.5, PDNC: 43.9 ± 7.6; p < 0.01). Additionally, the PDCI group showed more severe motor symptoms than the PDNC group (MDS-UPDRS Part III: 24.0 ± 9.7 vs 18.1 ± 7.1, p = 0.0074).

**Table 2.**
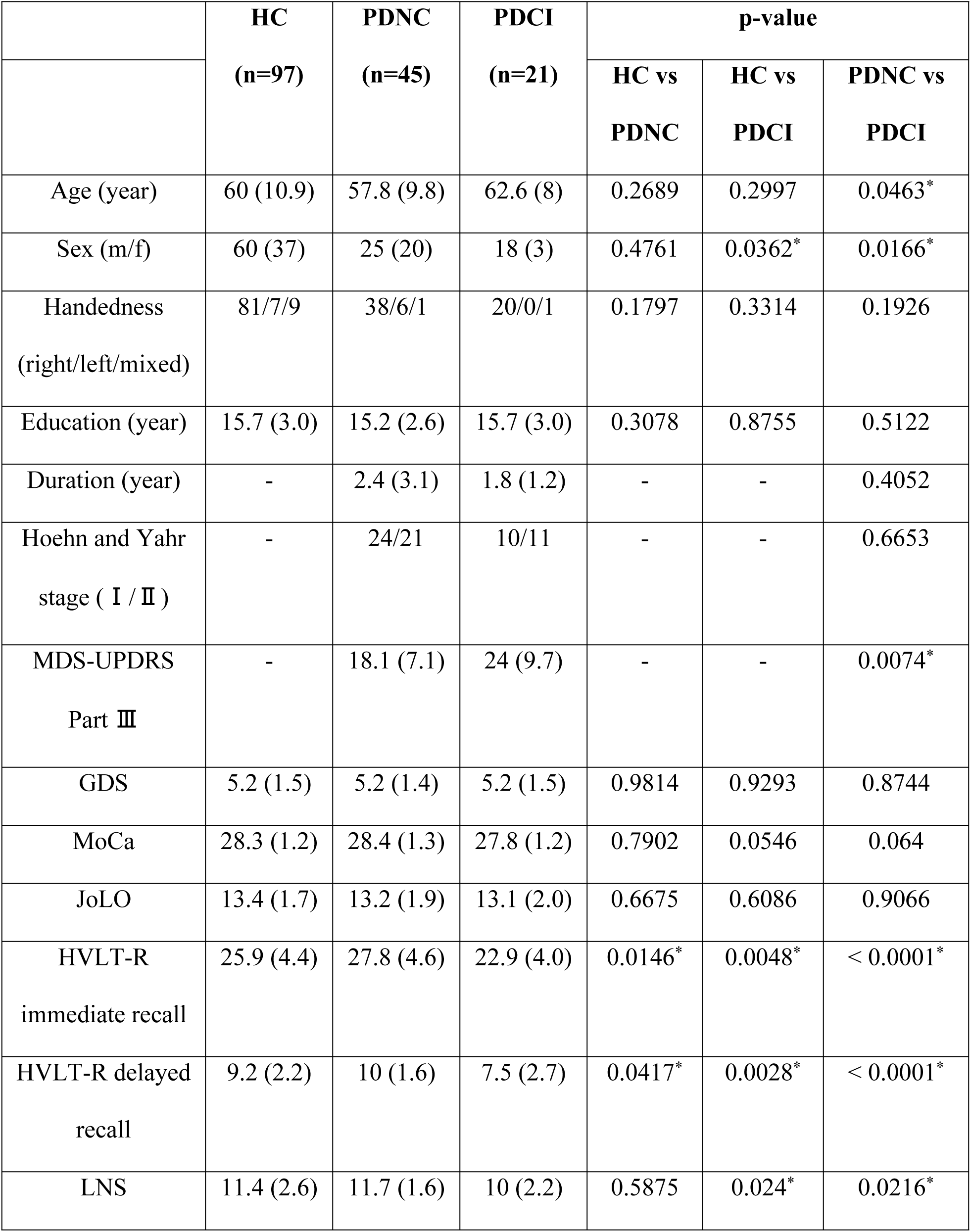
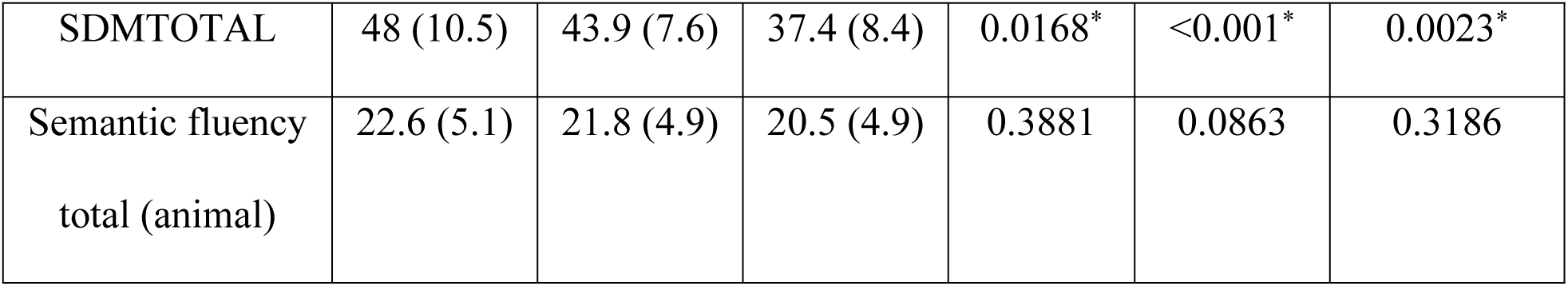
Demographic and clinical characteristics. HC: Healthy control, PDNC: Parkinson’s disease with normal cognition, PDCI: Parkinson’s Disease patients who show cognitive impairment, MDS-UPDRS: Movement Disorder Society– Unified Parkinson’s Disease Rating Scale, GDS: Geriatric Depression Scale, MoCA: Montreal Cognitive Assessment, JoLO: Judgment of Line Orientation, HVLT-R: Hopkins Verbal Learning Test-Revised, LNS: Letter-Number Sequencing, SDMT: Symbol Digit Modalities Test.

### Comparison of Intracranial Occupancy Rates

Table 3 shows significant differences in specific subregions of the amygdala, thalamus, hypothalamus, and hippocampus. In the amygdala, the PDCI group showed significantly lower intracranial occupancy rates in the left basal nucleus than the HC group. In the thalamus, the PDCI group showed significantly lower intracranial occupancy rates in the right LGN, right MDm, right PuA, and left LGN than the HC group. Furthermore, the PDCI group showed significantly lower intracranial occupancy rates in the left LGN than the PDNC group. In the hypothalamus, the PDCI group showed significantly lower intracranial occupancy rates in the right anterior-superior, left anterior-superior, left superior tubular, and left total than the HC group.

**Table 3.**
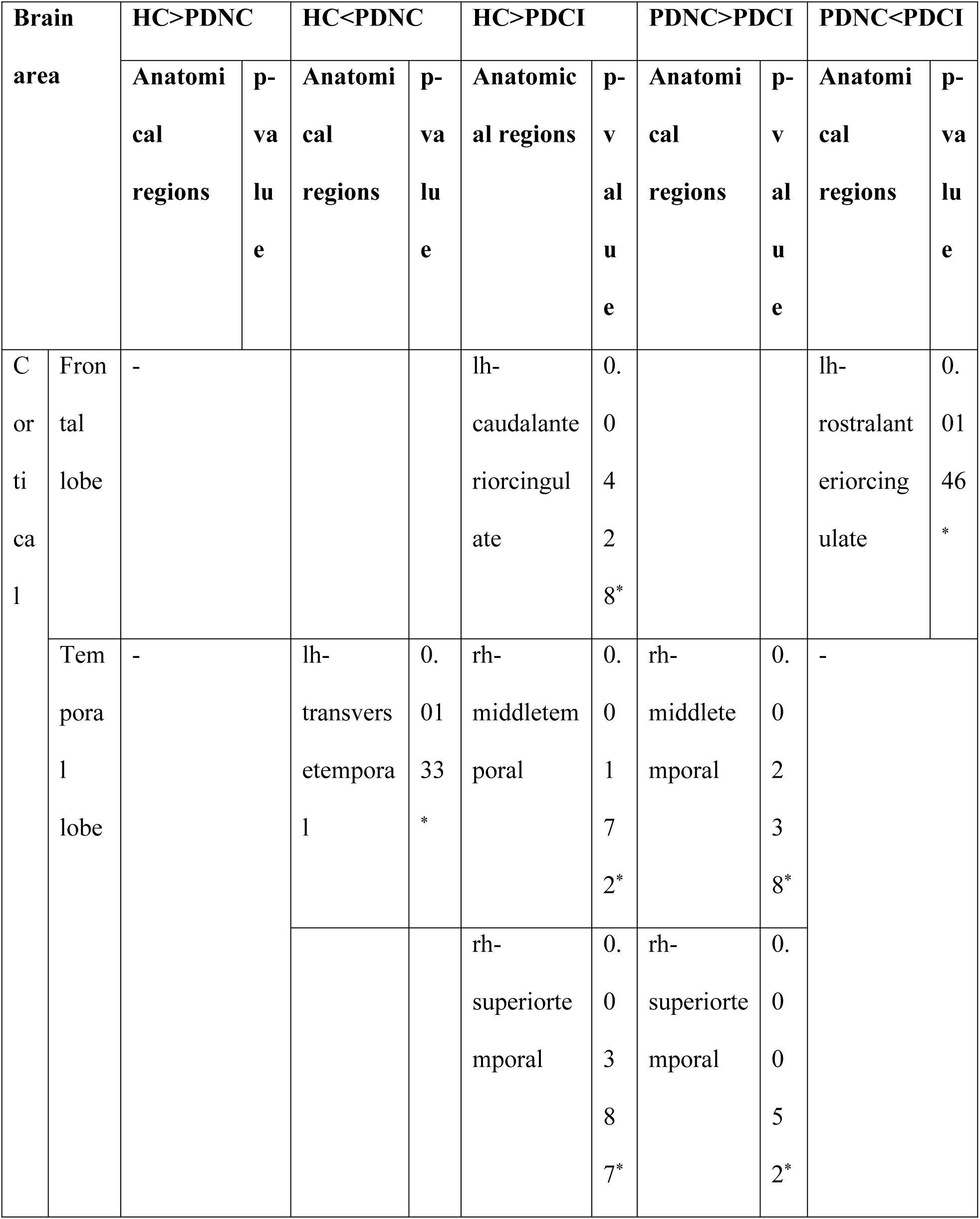
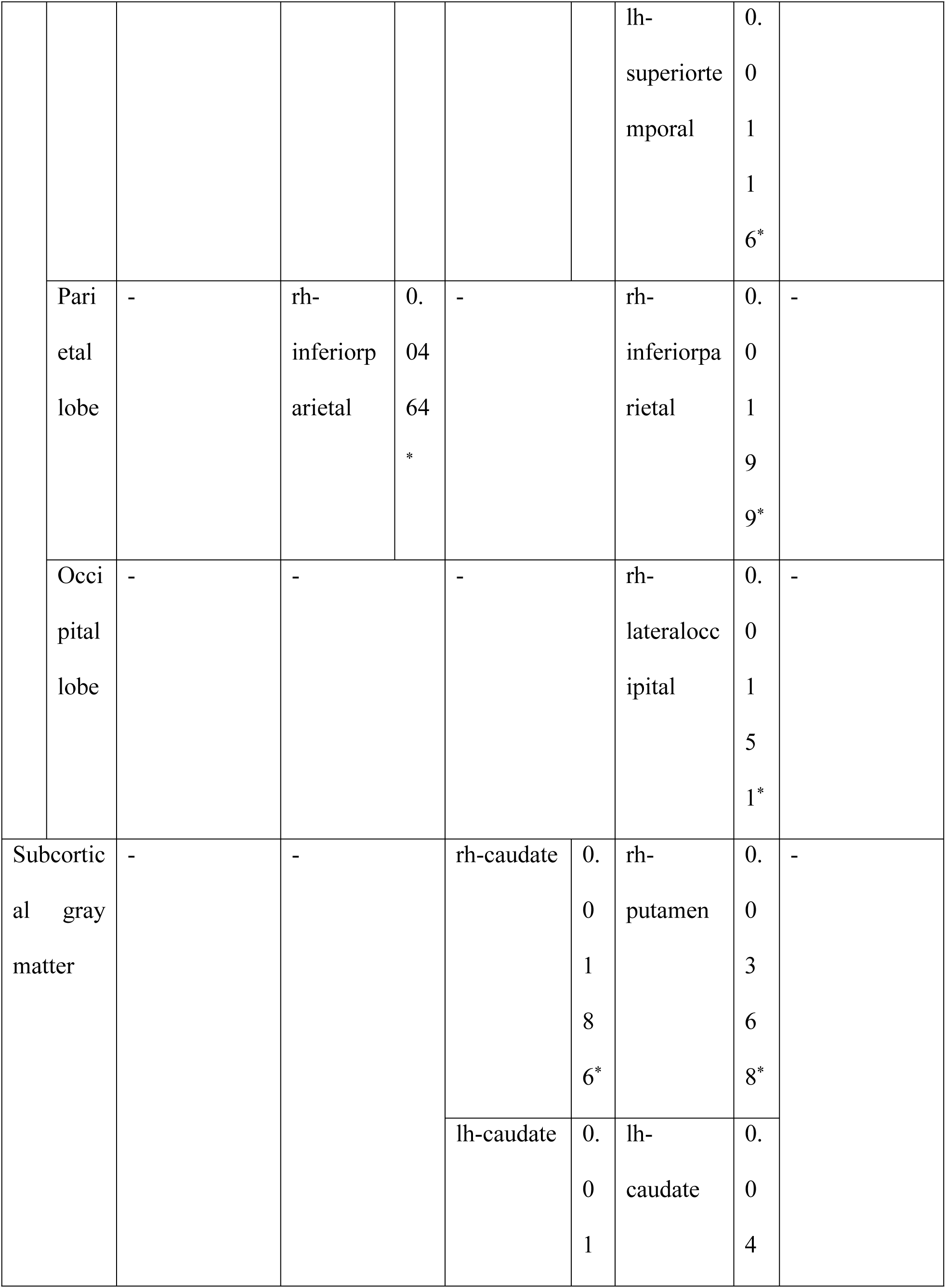
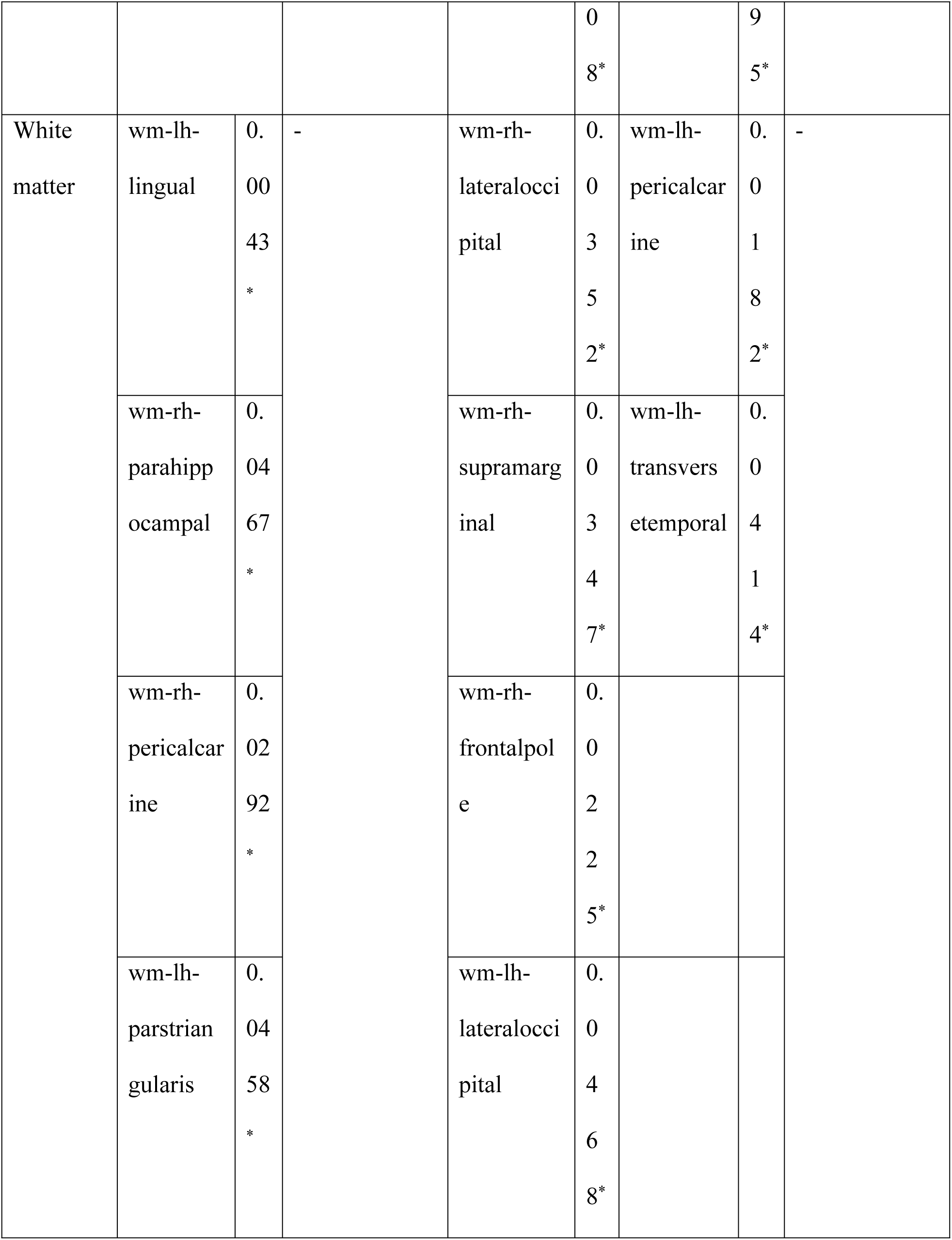
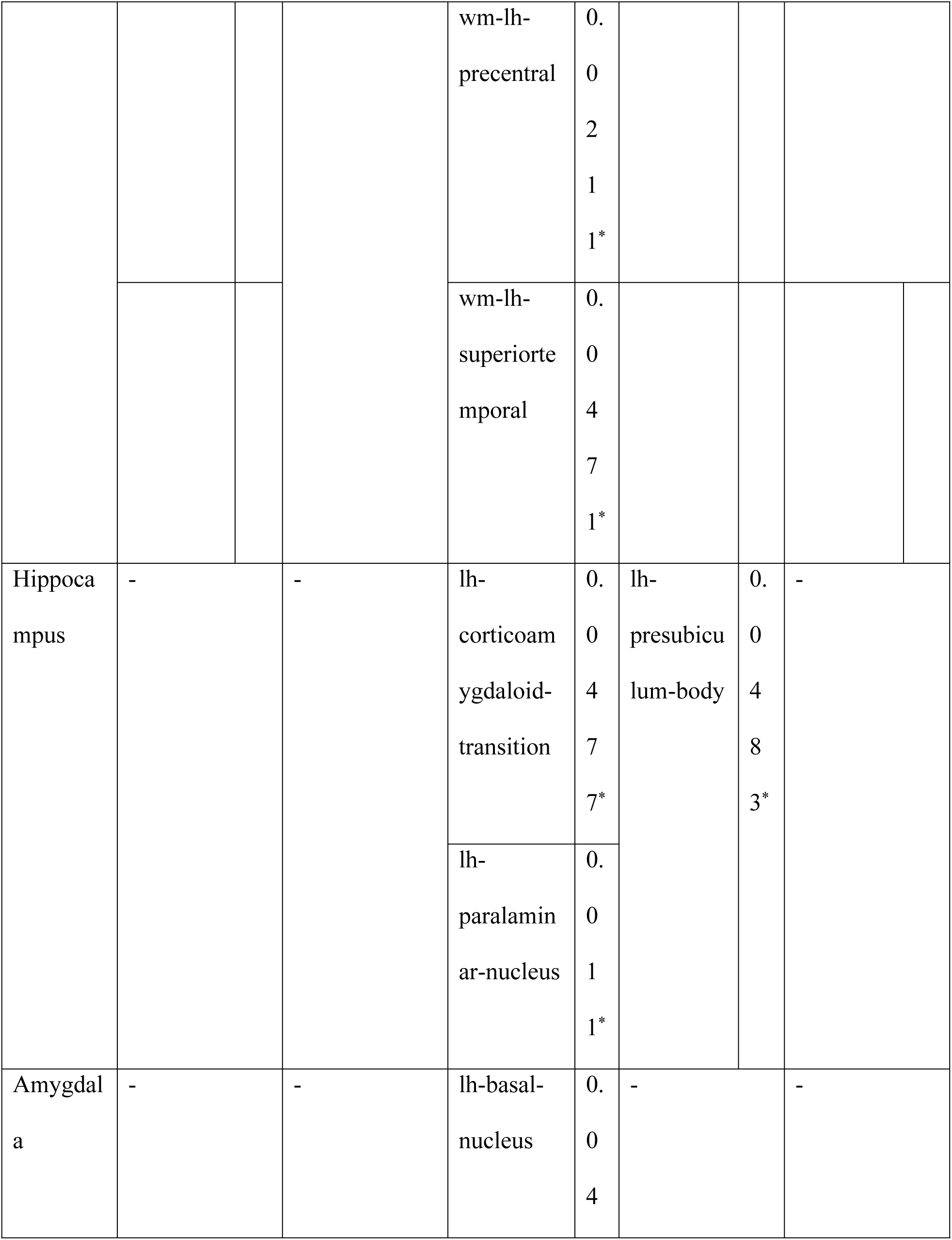
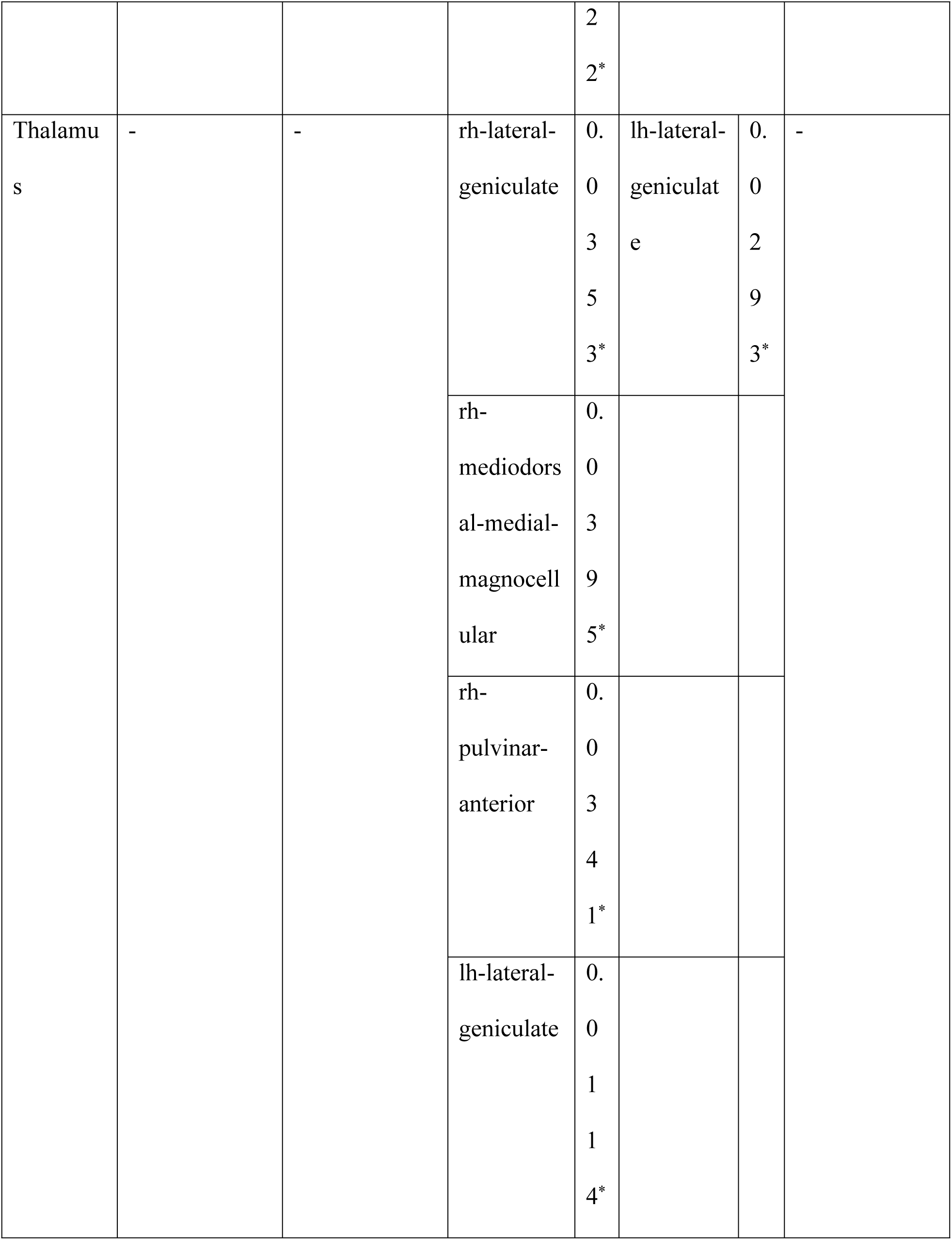
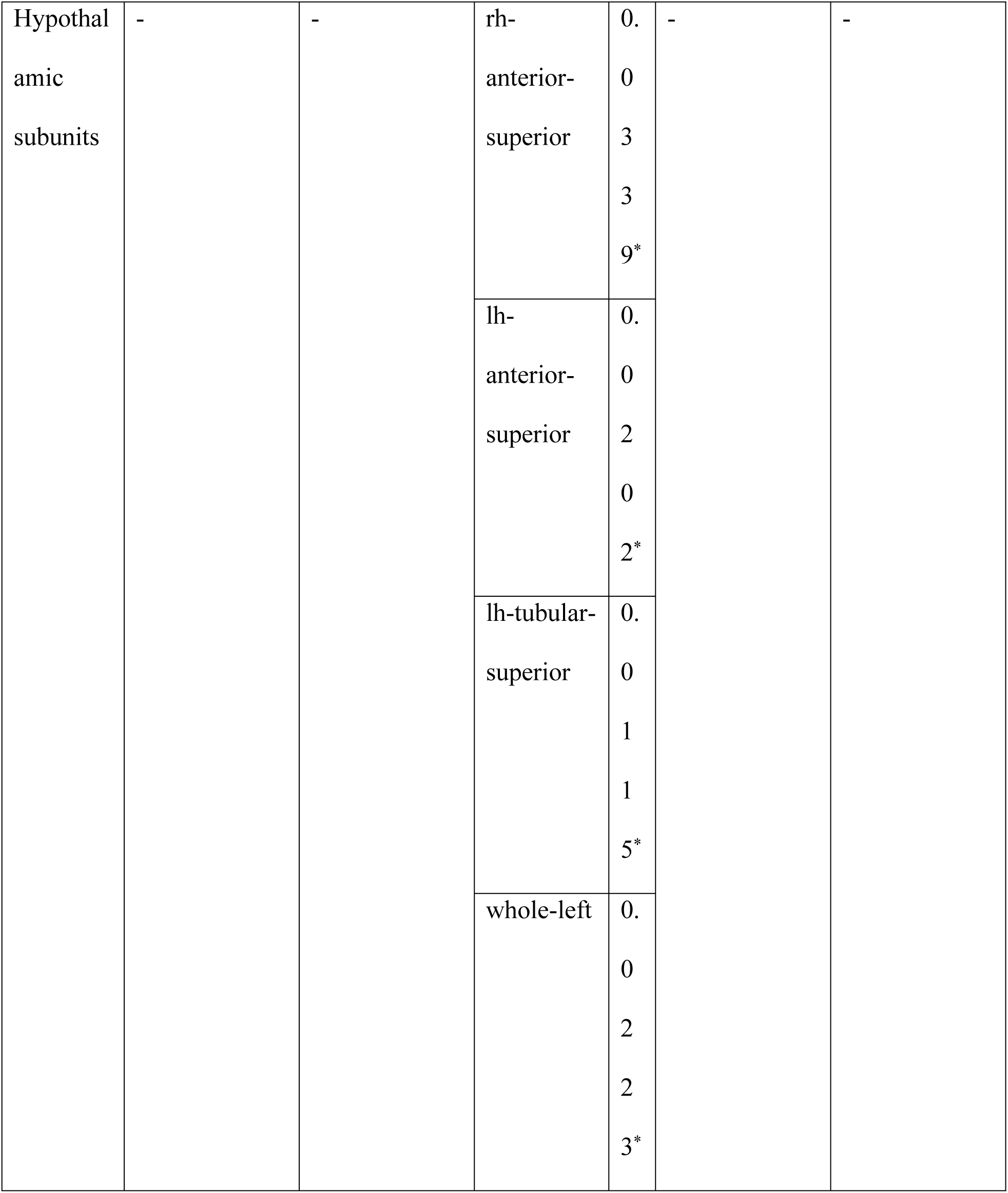
Between-group comparison of intracranial occupancy rates. HC: Healthy control, PDNC: Parkinson’s disease with normal cognition, PDCI: Parkinson’s disease patients who show cognitive impairment.

In hippocampal subregions, the PDCI group showed significantly lower intracranial occupancy rates in the left corticoamygdaloid transition area and left paralaminar nucleus than the HC group. The PDCI group also showed significantly lower intracranial occupancy rates in the left hippocampal parasubiculum-body than the PDNC group.

Significant differences were also observed in other brain regions. The PDCI group showed significantly lower intracranial occupancy rates than the HC group in the frontal lobe (left caudal anterior cingulate), temporal lobe (right middle temporal gyrus, right superior temporal gyrus), subcortical gray matter (right caudate nucleus, left caudate nucleus), and white matter regions (right lateral occipital, right supramarginal gyrus, right frontal pole, left lateral occipital, left precentral gyrus, left superior temporal).

The PDCI group showed significantly lower intracranial occupancy rates than the PDNC group in the temporal lobe (right middle temporal gyrus, right superior temporal gyrus, left superior temporal gyrus), parietal lobe (right inferior parietal), subcortical gray matter (left caudate nucleus, right putamen), and white matter regions (right lateral occipital, left transverse temporal, left pericalcarine). Conversely, the PDCI group showed significantly higher intracranial occupancy rates in the left rostral anterior cingulate than the PDNC group.

The PDNC group showed significantly lower intracranial occupancy rates than the HC group in white matter regions (left lingual gyrus, right parahippocampal gyrus, right pericalcarine, left pars triangularis of inferior frontal gyrus). However, the PDNC group showed significantly higher intracranial occupancy rates in the left transverse temporal gyrus and right inferior parietal than the HC group.

### Association Between Cognitive Test Scores and Brain Regions

As shown in Table 4, we analyzed the association between cognitive test scores and brain regions that showed significant differences in intracranial occupancy rates between PDCI and PDNC groups. No significant associations were found for subregions of the amygdala, thalamus, and hypothalamus.

**Table 4.**
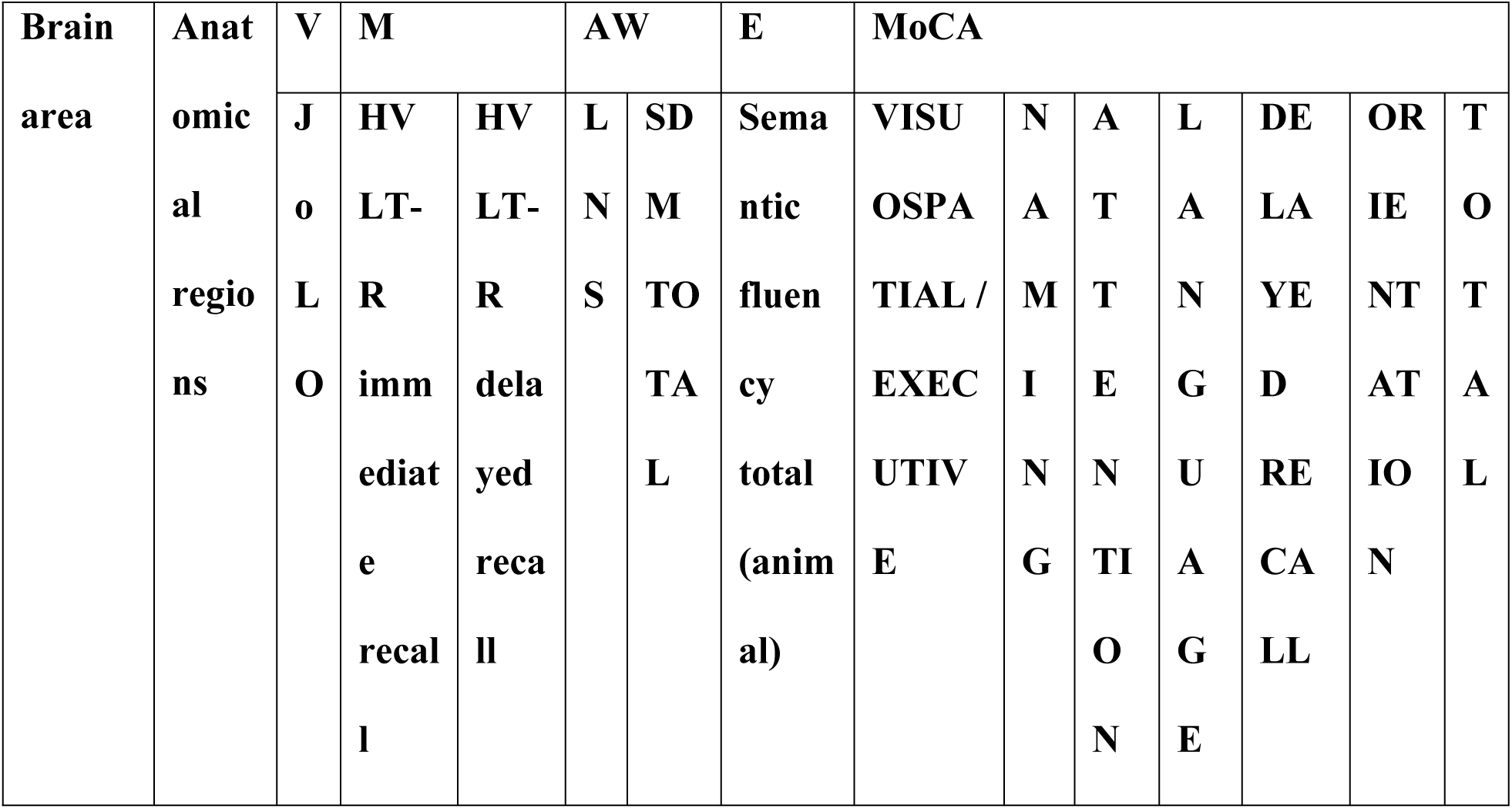
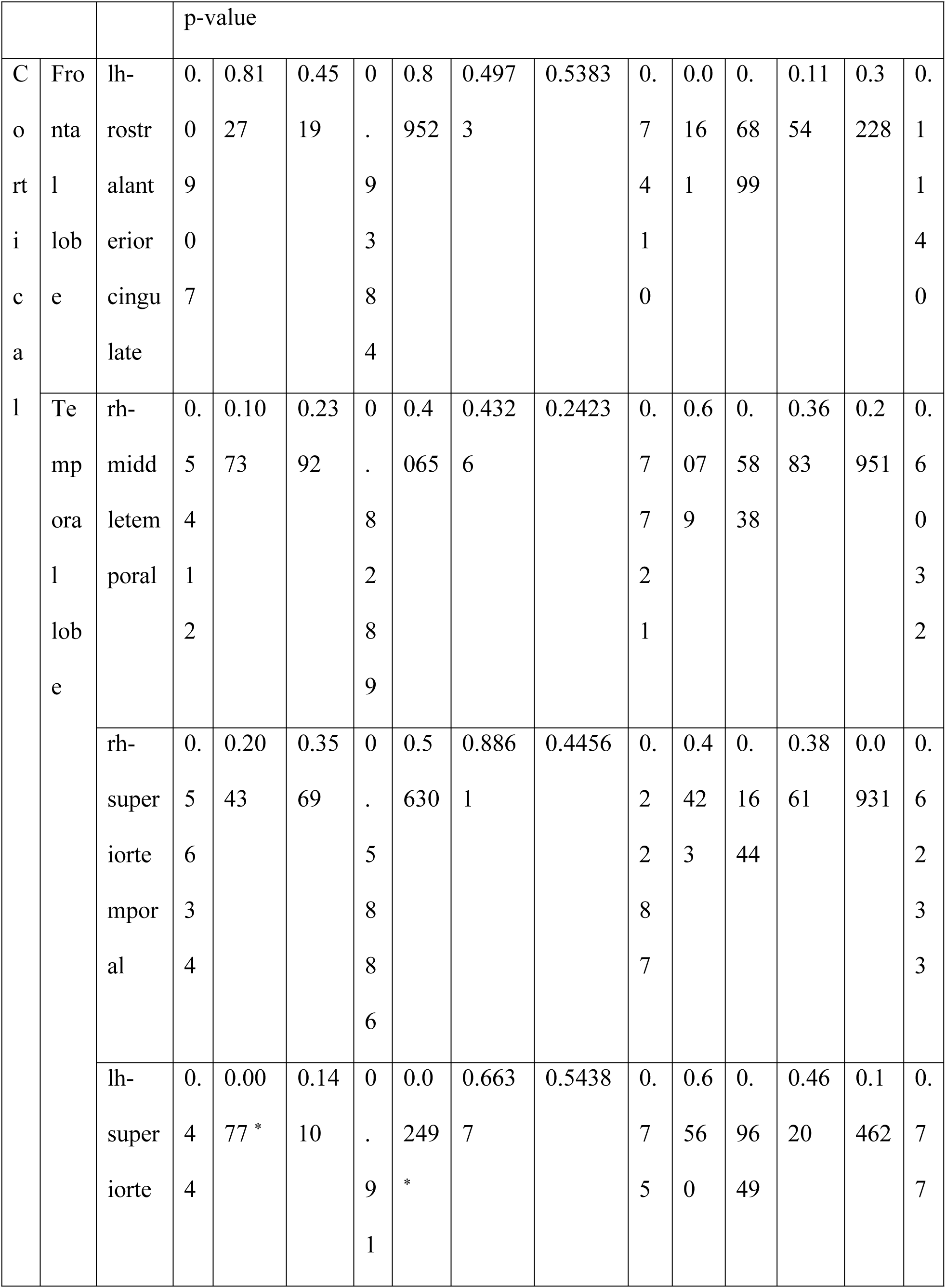
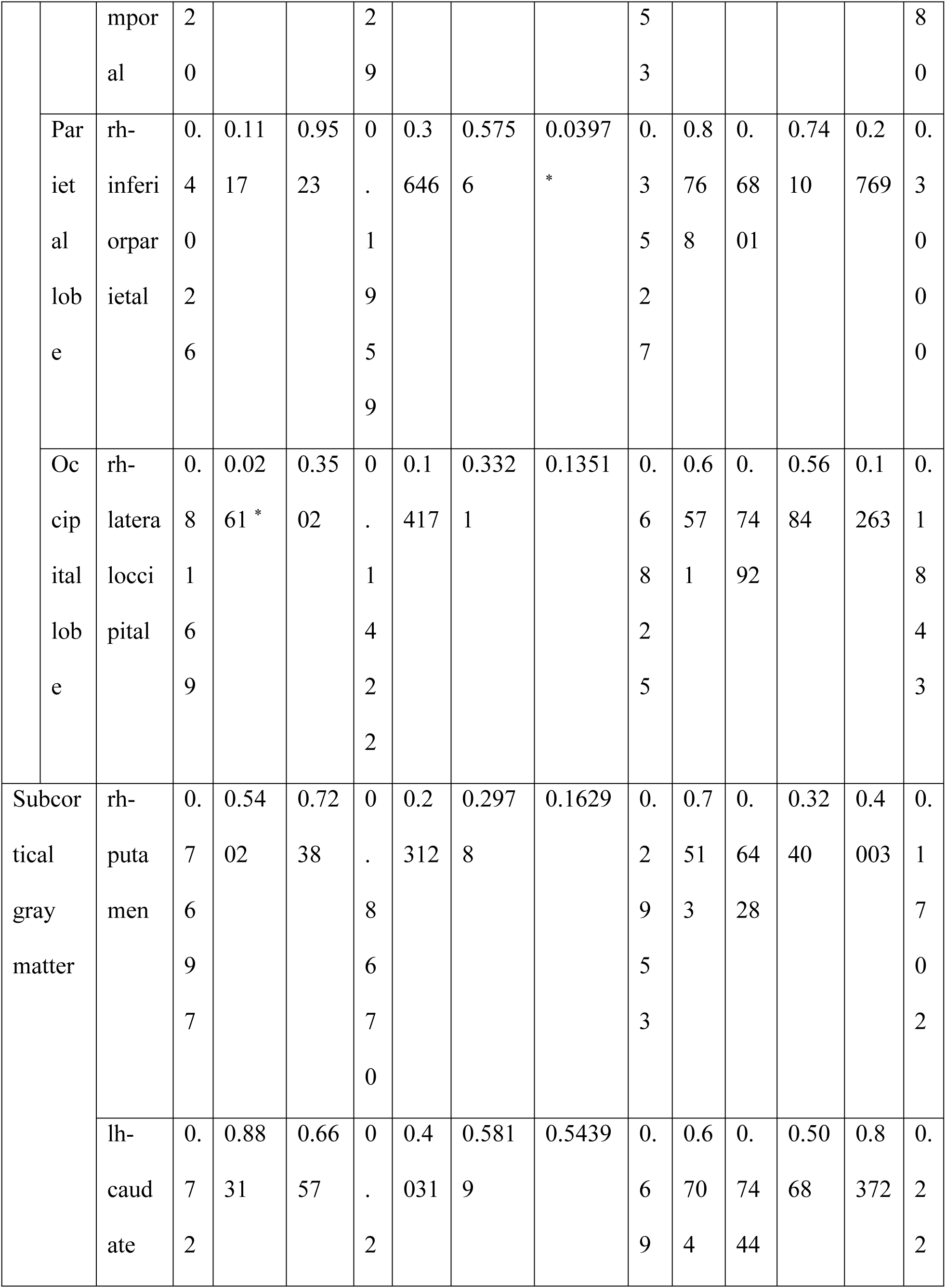
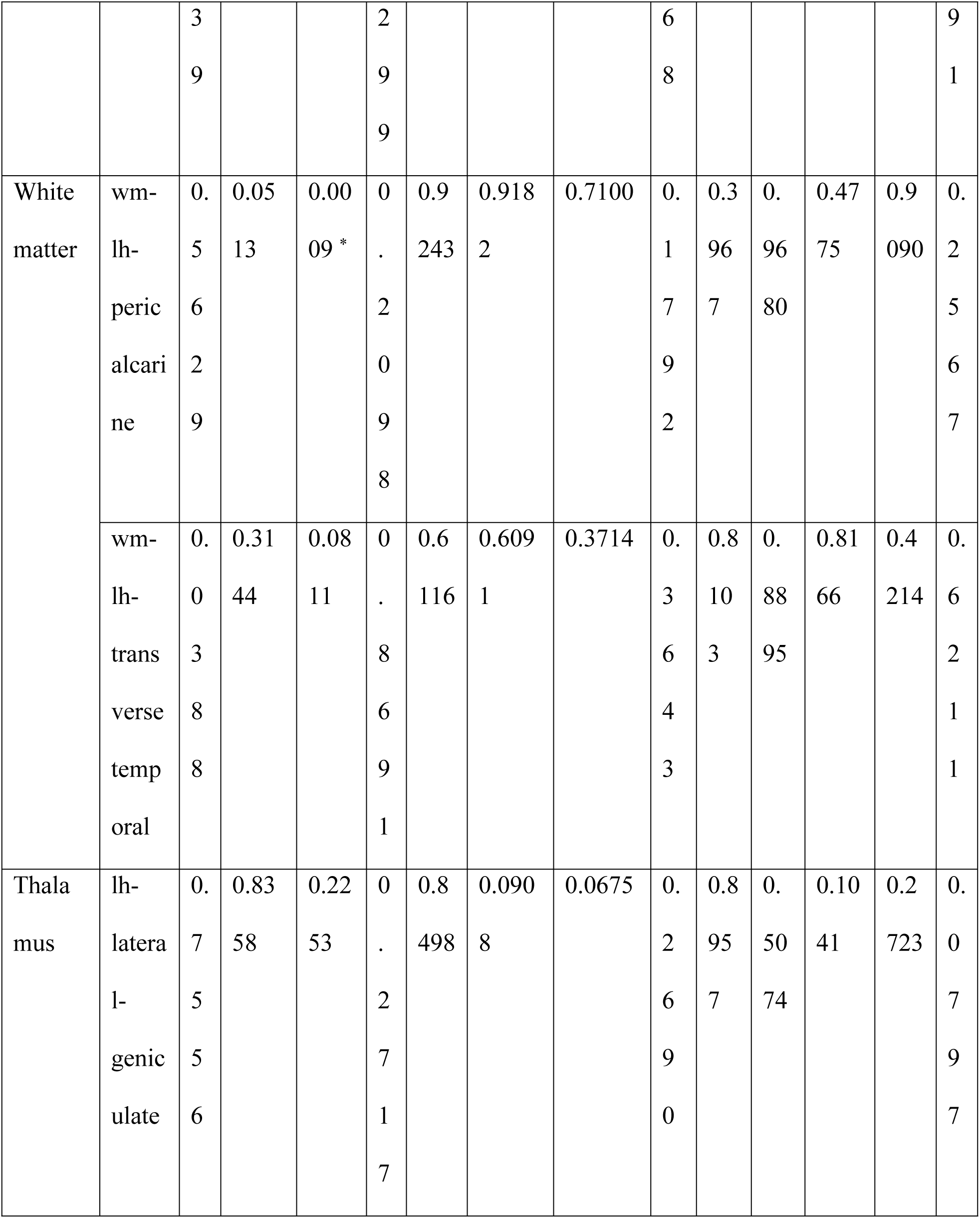
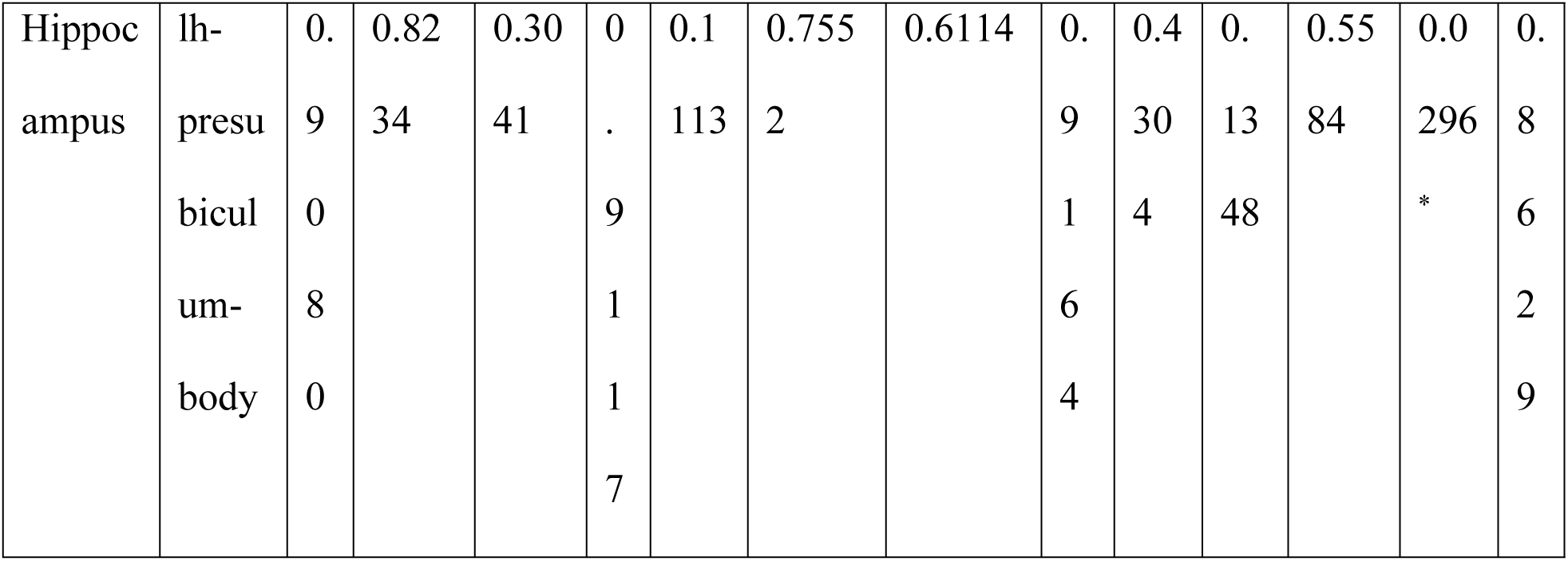
Association with cognitive test scores. PDNC: Parkinson’s disease with normal cognition, PDCI: Parkinson’s disease patients who show cognitive impairment, JoLO: Judgment of Line Orientation, HVLT-R: Hopkins Verbal Learning Test-Revised, LNS: Letter-Number Sequencing, SDMTOTAL: Total score of Symbol Digit Modalities Test, MoCA: Montreal Cognitive Assessment.

In hippocampal subregions, the left hippocampal parasubiculum-body showed a significant association with the orientation component of MoCA (p = 0.0296). In other brain regions, the left superior temporal gyrus was significantly associated with HVLT-R immediate recall (p = 0.0077) and SDMTOTAL (p = 0.0249), the right lateral occipital region with HVLT-R immediate recall (p = 0.0261), the left transverse temporal white matter region with JoLO score (p = 0.0388), the right inferior parietal region with the visuospatial/executive function component of MoCA (p = 0.0397), the left rostral anterior cingulate with the attention component of MoCA (p = 0.0161), and the left transverse temporal white matter with HVLT-R delayed recall (p = 0.0009).

### Hazard Ratios for MCI Progression

Table 5 shows the hazard ratios for MCI progression calculated based on the intracranial occupancy rates of brain regions that showed significant differences between PDCI and PDNC groups. However, no significant hazard ratios were observed in the analyzed brain regions.

**Table 5.** Hazard ratios for MCI progression in brain regions with significant differences between PDCI and PDNC. MCI: mild cognitive impairment, PDNC: Parkinson’s disease with normal cognition, PDCI: Parkinson’s disease patients who show cognitive impairment.

| Anatomical regions | Hazard ratio per 1 SD | p-value |
| --- | --- | --- |
| lh-rostralanteriorcingulate | 1.22 | 0.1115 |
| rh-middletemporal | 1.20 | 0.2880 |
| rh-superiortemporal | 1.28 | 0.1054 |
| lh-superiortemporal | 1.25 | 0.1186 |
| rh-inferiorparietal | 1.24 | 0.1402 |
| rh-lateraloccipital | 1.21 | 0.1510 |
| rh-putamen | 1.17 | 0.2617 |
| lh-caudate | 1.12 | 0.3883 |
| wm-lh-pericalcarine | 1.20 | 0.1843 |
| wm-lh-transversetemporal | 1.19 | 0.2213 |
| lh-lateral-geniculate | 1.20 | 0.1826 |
| lh-presubiculum-body | 1.79 | 0.3370 |

## Discussion

This study provides important insights into early structural brain changes associated with cognitive decline in PD through detailed subregional analysis of subcortical structures, enabled by recent advances in MRI technology. Our analysis revealed specific structural alterations in the amygdala (left basal nucleus), thalamus (bilateral lateral geniculate nucleus), and hypothalamus (anterior-superior region) in PDCI patients compared with HC—findings that have not been previously reported at this level of anatomical detail.

From a morphological perspective, widespread structural changes were observed in cortical areas, subcortical gray matter, white matter regions, and hippocampal subregions. Interestingly, while volume changes in the amygdala, thalamus, and hypothalamus subregions did not directly correlate with cognitive test scores, structural changes in various cortical regions were associated with specific cognitive functions. These findings suggest a complex relationship between brain structure and function in PD-related cognitive decline.

Hazard ratio analysis for MCI progression did not yield significant results for individual brain regions. This suggests that cognitive decline in PD is likely caused by changes across multiple regions and overall network alterations, rather than changes in specific brain areas.

On comparing PDCI and HC, we detected subtle structural abnormalities in subregions of the amygdala, thalamus, and hypothalamus. The changes observed in the amygdala suggest its importance in “brain-first” type PD. Previous studies have indicated that in brain-first type PD, the amygdala shows pathological changes from early stages, associated with early cognitive dysfunction [33, 34]. Lewy body accumulation in the amygdala may serve as a starting point for pathological expansion to other brain regions, potentially leading to cognitive and emotional dysfunction. Notably, previous research has shown a concentration of Lewy bodies in the basolateral nucleus of the amygdala [35]. In the thalamus, MRI studies have confirmed iron accumulation, suggesting that this abnormality aligns with the distribution of Lewy body pathology and is closely related to cognitive decline [36]. Contrastingly, DTI studies have shown marked deterioration in the microstructure of projection fibers associated with specific thalamic nuclei, but the fine regions detected in our study have not been evaluated [37]. Furthermore, arterial spin labeling studies have reported that decreased blood flow in the dorsomedial nucleus (MDm) of the thalamus is closely related to cognitive decline [38]. Additionally, a significant reduction in overall thalamic volume has been observed in PD patients, which may be an early phenomenon associated with disease progression [39]. Functional MRI studies have shown how hypothalamic degeneration is related to autonomic dysfunction in PD patients [40]. Hypothalamic degeneration may indirectly affect cognitive function through autonomic nervous system and metabolic regulation [32].

Besides these subcortical structures, the PDCI group also showed structural abnormalities in cortical areas (frontal and temporal lobes), subcortical gray matter (caudate nucleus), white matter regions, and hippocampal subregions. These results suggest that cognitive decline in PD is associated with widespread structural changes across the brain, rather than localized changes.

Interestingly, the comparison between PDCI and PDNC showed no significant differences in the amygdala or hypothalamus regions. However, significant differences were observed in cortical areas (temporal and parietal lobes), subcortical gray matter (caudate nucleus, putamen), white matter regions, hippocampal subregions, and some thalamic subregions (left LGN). The lack of significant differences in the amygdala may be owing to the amygdala being affected early in the pathological process, suggesting that some changes have already occurred at the PDNC stage. However, contrary to expectations, no significant differences were observed in the amygdala between PDNC and HC. This result suggests that structural changes in the amygdala in the early stages of PD are complex and involve compensatory changes [41, 42] or other factors beyond simple atrophy.

Our study confirmed that structural changes in various brain regions in PD patients are closely related to specific cognitive functions. Our study confirmed that structural changes in various brain regions in PD patients are closely related to specific cognitive functions. The left superior temporal gyrus is associated with a decline in memory and attention/processing speed. MRI studies have shown that decreased cortical thickness in this region is related to reduced memory function [43]. Furthermore, magnetoencephalography studies have revealed that decreased functional connectivity between the left superior temporal gyrus and other brain regions is associated with reduced attention and processing speed [44]. The right lateral occipital region’s gray matter volume reduction is associated with physical frailty and cognitive impairment [45], while decreased glucose metabolism in this area is closely related to memory function decline [46]. Degeneration in the left transverse temporal white matter region has been linked to decreased visuospatial function, indicating that white matter degeneration affects cognitive function in PD patients [47]. Regarding the right inferior parietal region, gray matter reduction and decreased functional connectivity are associated with a decline in visuospatial and executive functions [48, 49]. The left hippocampal parasubiculum volume reduction is associated with cognitive decline and is an important factor in predicting the risk of transition to dementia [16]. The left rostral anterior cingulate is linked to decreased attention function, with observed reduced blood flow during attention-demanding sentence processing tasks [50]. These findings highlight the complex relationship between localized brain structural changes and specific cognitive impairments in PD.

In our hazard ratio analysis, we calculated the hazard ratios for MCI progression for brain regions that showed significant differences between PDCI and PDNC. However, no significant results were obtained for individual brain regions. This result suggests that cognitive decline in PD is caused by changes in multiple regions and overall network changes, rather than changes in a single brain region [51, 52]. This finding represents a key conclusion of our study, highlighting the complex, interconnected nature of brain changes associated with cognitive decline in PD. It also suggests that a more comprehensive approach is needed to predict the risk of cognitive decline, rather than just focusing on changes in individual brain regions [53–57]. Future research should aim to develop predictive models that incorporate multiple brain regions and their interactions to more accurately assess cognitive decline risk in PD patients.

A key finding of this study is that despite detecting subtle structural changes in the amygdala, thalamus, and hypothalamus, no direct correlation was found between volume changes in these regions and cognitive test scores. Conversely, widespread cortical areas showed early impairment, and these changes were associated with cognitive function. These results may seem to contradict the traditional progression model of Lewy body pathology.

The mechanisms of cortical impairment and cognitive decline in PD are suggested to be a complex process. This mechanism is thought to involve the interaction of multiple systems, including pathological changes [56], network-level degeneration [4], and compensatory adaptation [57, 58], among others [59]. Notably, this complex interaction suggests that abnormalities in neural circuits are already causing cognitive impairment before Lewy body pathology clearly progresses to the cortex [60].

Therefore, the importance of early diagnosis and intervention has been emphasized. For example, interventions targeting specific brain regions or approaches aimed at improving function at the network level could be considered. These may include pharmacotherapy targeting specific neural circuits, non-invasive brain stimulation therapy, or cognitive function training. Future studies should focus on these aspects.

One limitation of this study is the relatively small sample size. This is mainly owing to the extraction of only subjects with identical MRI protocols to accurately analyze fine subregions. While this strict selection criterion enhances the precision of image analysis, it limited the sample size.

Additionally, our use of MoCA as the primary criterion for MCI determination warrants discussion. While the MDS Task Force guidelines suggest a more comprehensive Level II diagnostic approach using multiple cognitive domain tests, we employed MoCA (Level I) based on several considerations. MoCA has been validated as an effective tool for assessing cognitive function in PD patients and has demonstrated reliability in predicting cognitive decline. Studies have shown comparable effectiveness between Level I and Level II criteria in identifying PD-MCI, and MoCA has proven particularly useful for predicting long-term cognitive outcomes. Furthermore, this approach aligns with real-world clinical practice, where time constraints often necessitate the use of screening tools rather than extensive neuropsychological batteries.

Nevertheless, we acknowledge that a more comprehensive neuropsychological assessment might have provided deeper insights into domain-specific cognitive impairments. Our baseline data revealed that the PDCI group showed lower performance across several cognitive domains than the PDNC group, suggesting that subtle cognitive changes might be detectable through detailed domain-specific testing. Future research combining both screening tools and comprehensive neuropsychological assessments could better illuminate the relationship between specific cognitive domain impairments and structural brain changes in early PD. Despite these limitations, this study provides important insights into the relationship between early brain structural changes and cognitive decline in PD.

In conclusion, this study highlights early structural brain changes linked to cognitive decline in PD, with specific alterations in the amygdala, thalamus, and hypothalamus subregions in PDCI patients compared with HCs. While widespread structural changes were noted in cortical and subcortical areas, only cortical alterations were associated with specific cognitive functions, indicating that cognitive decline in PD results from widespread network alterations rather than changes in specific regions. These insights could enhance our understanding of PD-related cognitive decline and inform early diagnostic and therapeutic strategies. Future research should develop predictive models that integrate multiple brain regions and their interactions to further unravel mechanisms underlying cognitive decline in PD.

## Data Availability

Data used in the preparation of this article were obtained [on Feb, 23 2023] from the Parkinson's Progression Markers Initiative (PPMI) database (www.ppmi-info.org/access-dataspecimens/download-data), RRID:SCR 006431. Access to the PPMI database requires user registration at the PPMI website. For up-to-date information on the study, visit www.ppmi-info.org

## Acknowledgment

We acknowledge the use of the artificial intelligence (AI) language model, Claude 3.5 Sonnet, developed by Anthropic, for assistance with manuscript editing and English language proofreading. This AI tool was employed to enhance the clarity and readability of our manuscript. However, all scientific content, analysis, and conclusions were generated and verified by the human authors. The use of this AI tool does not alter the scientific integrity of our work, and we maintain full responsibility for the content of this paper.

Data used in the preparation of this article were obtained [on Feb, 23 2023] from the Parkinson’s Progression Markers Initiative (PPMI) database (www.ppmi-info.org/access-dataspecimens/download-data), RRID:SCR 006431. For up-to-date information on the study, visit www.ppmi-info.org “PPMI – a public-private partnership – is funded by the Michael J. Fox Foundation for Parkinson’s Research and funding partners, including [list the full names of all of the PPMI funding partners found on the PPMI Website].”

## Notes

### Competing Interest Statement

The authors have declared no competing interest.

### Funding Statement

The author(s) received no specific funding for this work.

### Author Declarations

The PPMI is registered at the National Institutes of Health’s clinical trial registry (NCT04477785).

## References

1. Kalia LV, Lang AE. Parkinson’s disease. Lancet. 2015;386(9996): 896–912.

2. Leroi I, McDonald K, Pantula H, Harbishettar V. Cognitive impairment in Parkinson disease: impact on quality of life, disability, and caregiver burden. J Geriatr Psychiatry Neurol. 2012;25(4): 208–214.

3. Litvan I, Goldman JG, Troster AI, Schmand BA, Weintraub D, Petersen RC, et al. Diagnostic criteria for mild cognitive impairment in Parkinson’s disease: Movement Disorder Society Task Force guidelines. Mov Disord. 2012;27(3): 349–356.

4. Gratwicke J, Jahanshahi M, Foltynie T. Parkinson’s disease dementia: a neural networks perspective. Brain. 2015;138(Pt 6): 1454–1476.

5. Compta Y, Parkkinen L, O’Sullivan SS, Vandrovcova J, Holton JL, Collins C, et al. Lewy- and Alzheimer-type pathologies in Parkinson’s disease dementia: which is more important? Brain. 2011;134(Pt 5): 1493–1505.

6. Lerch JP, van der Kouwe AJ, Raznahan A, Paus T, Johansen-Berg H, Miller KL, et al. Studying neuroanatomy using MRI. Nat Neurosci. 2017;20(3): 314–326.

7. Lanskey JH, McColgan P, Schrag AE, Acosta-Cabronero J, Rees G, Morris HR, et al. Can neuroimaging predict dementia in Parkinson’s disease? Brain. 2018;141(9): 2545–2460.

8. Hou Y, Shang H. Magnetic resonance imaging markers for cognitive impairment in Parkinson’s disease: current view. Front Aging Neurosci. 2022;14: 788846.

9. Zheng D, Chen C, Song W, Yi Z, Zhao P, Zhong J, et al. Regional gray matter reductions associated with mild cognitive impairment in Parkinson’s disease: A meta-analysis of voxel-based morphometry studies. Behav Brain Res. 2019;371: 111973.

10. Mihaescu AS, Masellis M, Graff-Guerrero A, Kim J, Criaud M, Cho SS, et al. Brain degeneration in Parkinson’s disease patients with cognitive decline: a coordinate-based meta-analysis. Brain Imaging Behav. 2019;13(4): 1021–1034.

11. Xu Y, Yang J, Hu X, Shang H. Voxel-based meta-analysis of gray matter volume reductions associated with cognitive impairment in Parkinson’s disease. J Neurol. 2016;263(6): 1178–1187.

12. Mak E, Su L, Williams GB, Firbank MJ, Lawson RA, Yarnall AJ, et al. Baseline and longitudinal grey matter changes in newly diagnosed Parkinson’s disease: ICICLE-PD study. Brain. 2015;138(Pt 10): 2974–2986.

13. Foo H, Mak E, Yong TT, Wen MC, Chander RJ, Au WL, et al. Progression of subcortical atrophy in mild Parkinson’s disease and its impact on cognition. Eur J Neurol. 2017;24(2): 341–348.

14. Gorges M, Kunz MS, Muller HP, Liepelt-Scarfone I, Storch A, Dodel R, et al. Longitudinal brain atrophy distribution in advanced Parkinson’s disease: What makes the difference in “cognitive status” converters? Hum Brain Mapp. 2020;41(6): 1416–1434.

15. Filippi M, Canu E, Donzuso G, Stojkovic T, Basaia S, Stankovic I, et al. Tracking cortical changes throughout cognitive decline in Parkinson’s disease. Mov Disord. 2020;35(11): 1987–1998.

16. Foo H, Mak E, Chander RJ, Ng A, Au WL, Sitoh YY, et al. Associations of hippocampal subfields in the progression of cognitive decline related to Parkinson’s disease. Neuroimage Clin. 2017;14: 37–42.

17. Iglesias JE, Augustinack JC, Nguyen K, Player CM, Player A, Wright M, et al. A computational atlas of the hippocampal formation using ex vivo, ultra-high resolution MRI: Application to adaptive segmentation of in vivo MRI. Neuroimage. 2015;115: 117–137.

18. Saygin ZM, Kliemann D, Iglesias JE, van der Kouwe AJW, Boyd E, Reuter M, et al. High-resolution magnetic resonance imaging reveals nuclei of the human amygdala: manual segmentation to automatic atlas. Neuroimage. 2017;155: 370–382.

19. Iglesias JE, Insausti R, Lerma-Usabiaga G, Bocchetta M, Van Leemput K, Greve DN, et al. A probabilistic atlas of the human thalamic nuclei combining ex vivo MRI and histology. Neuroimage. 2018;183: 314–326.

20. Billot B, Bocchetta M, Todd E, Dalca AV, Rohrer JD, Iglesias JE. Automated segmentation of the hypothalamus and associated subunits in brain MRI. Neuroimage. 2020;223: 117287.

21. Becker S, Granert O, Timmers M, Pilotto A, Van Nueten L, Roeben B, et al. Association of hippocampal subfields, CSF biomarkers, and cognition in patients with Parkinson disease without dementia. Neurology. 2021;96(6): e904–e915.

22. Kandiah N, Zainal NH, Narasimhalu K, Chander RJ, Ng A, Mak E, et al. Hippocampal volume and white matter disease in the prediction of dementia in Parkinson’s disease. Parkinsonism Relat Disord. 2014;20(11): 1203–1208.

23. Braak H, Braak E, Yilmazer D, de Vos RA, Jansen EN, Bohl J, et al. Amygdala pathology in Parkinson’s disease. Acta Neuropathol. 1994;88(6): 493–500.

24. Huang P, Xuan M, Gu Q, Yu X, Xu X, Luo W, et al. Abnormal amygdala function in Parkinson’s disease patients and its relationship to depression. J Affect Disord. 2015;183: 263–268.

25. Ziabreva I, Ballard CG, Aarsland D, Larsen JP, McKeith IG, Perry RH, et al. Lewy body disease: thalamic cholinergic activity related to dementia and parkinsonism. Neurobiol Aging. 2006;27(3): 433–438.

26. Sandyk R, Iacono RP, Bamford CR. The hypothalamus in Parkinson disease. Ital J Neurol Sci. 1987;8(3): 227–234.

27. Dayan E, Sklerov M, Browner N. Disrupted hypothalamic functional connectivity in patients with PD and autonomic dysfunction. Neurology. 2018;90(23): e2051–e2058.

28. Kim JS, Oh YS, Lee KS, Kim YI, Yang DW, Goldstein DS. Association of cognitive dysfunction with neurocirculatory abnormalities in early Parkinson disease. Neurology. 2012;79(13): 1323–1331.

29. Dalrymple-Alford JC, MacAskill MR, Nakas CT, Livingston L, Graham C, Crucian GP, et al. The MoCA: well-suited screen for cognitive impairment in Parkinson disease. Neurology. 2010;75(19): 1717–1725.

30. Hoogland J, Boel JA, de Bie RMA, Schmand BA, Geskus RB, Dalrymple-Alford JC, et al. Risk of Parkinson’s disease dementia related to level I MDS PD-MCI. Mov Disord. 2019;34(3): 430–435.

31. Kandiah N, Zhang A, Cenina AR, Au WL, Nadkarni N, Tan LC. Montreal Cognitive Assessment for the screening and prediction of cognitive decline in early Parkinson’s disease. Parkinsonism Relat Disord. 2014;20(11): 1145–1148.

32. Schrag A, Siddiqui UF, Anastasiou Z, Weintraub D, Schott JM. Clinical variables and biomarkers in prediction of cognitive impairment in patients with newly diagnosed Parkinson’s disease: a cohort study. Lancet Neurol. 2017;16(1): 66–75.

33. Borghammer P, Just MK, Horsager J, Skjaerbaek C, Raunio A, Kok EH, et al. A postmortem study suggests a revision of the dual-hit hypothesis of Parkinson’s disease. NPJ Parkinsons Dis. 2022;8(1): 166.

34. Horsager J, Borghammer P. Brain-first vs. body-first Parkinson’s disease: An update on recent evidence. Parkinsonism Relat Disord. 2024;122: 106101.

35. Harding AJ, Stimson E, Henderson JM, Halliday GM. Clinical correlates of selective pathology in the amygdala of patients with Parkinson’s disease. Brain. 2002;125(Pt 11): 2431–2445.

36. Thomas GEC, Leyland LA, Schrag AE, Lees AJ, Acosta-Cabronero J, Weil RS. Brain iron deposition is linked with cognitive severity in Parkinson’s disease. J Neurol Neurosurg Psychiatry. 2020;91(4): 418–425.

37. Planetta PJ, Schulze ET, Geary EK, Corcos DM, Goldman JG, Little DM, et al. Thalamic projection fiber integrity in de novo Parkinson disease. AJNR Am J Neuroradiol. 2013;34(1): 74–79.

38. Azamat S, Betul Arslan D, Erdogdu E, Kicik A, Cengiz S, Eryurek K, et al. dementia. Eur J Radiol. 2021;144: 109985.

39. Lee SH, Kim SS, Tae WS, Lee SY, Choi JW, Koh SB, et al. Regional volume analysis of the Parkinson disease brain in early disease stage: gray matter, white matter, striatum, and thalamus. AJNR Am J Neuroradiol. 2011;32(4): 682–687.

40. Dayan E, Sklerov M. Autonomic disorders in Parkinson disease: Disrupted hypothalamic connectivity as revealed from resting-state functional magnetic resonance imaging. Handb Clin Neurol. 2021;182: 211–222.

41. Blesa J, Trigo-Damas I, Dileone M, Del Rey NL, Hernandez LF, Obeso JA. Compensatory mechanisms in Parkinson’s disease: Circuits adaptations and role in disease modification. Exp Neurol. 2017;298(Pt B): 148–161.

42. Sanjari Moghaddam H, Dolatshahi M, Mohebi F, Aarabi MH. Structural white matter alterations as compensatory mechanisms in Parkinson’s disease: A systematic review of diffusion tensor imaging studies. J Neurosci Res. 2020;98(7): 1398–1416.

43. Lee EY. Memory deficits in Parkinson’s disease are associated with impaired attentional filtering and memory consolidation processes. J Clin Med. 2023;12(14).

44. Wiesman AI, Heinrichs-Graham E, McDermott TJ, Santamaria PM, Gendelman HE, Wilson TW. Quiet connections: Reduced fronto-temporal connectivity in nondemented Parkinson’s Disease during working memory encoding. Hum Brain Mapp. 2016;37(9): 3224–3235.

45. Chen YS, Chen HL, Lu CH, Chen MH, Chou KH, Tsai NW, et al. Reduced lateral occipital gray matter volume is associated with physical frailty and cognitive impairment in Parkinson’s disease. Eur Radiol. 2019;29(5): 2659–2668.

46. Zhihui S, Yinghua L, Hongguang Z, Yuyin D, Xiaoxiao D, Lulu G, et al. Correlation analysis between (18)F-fluorodeoxyglucose positron emission tomography and cognitive function in first diagnosed Parkinson’s disease patients. Front Neurol. 2023;14: 1195576.

47. Auning E, Kjaervik VK, Selnes P, Aarsland D, Haram A, Bjornerud A, et al. White matter integrity and cognition in Parkinson’s disease: a cross-sectional study. BMJ Open. 2014;4(1): e003976.

48. Zhang L, Wang M, Sterling NW, Lee EY, Eslinger PJ, Wagner D, et al. Cortical Thinning and Cognitive Impairment in Parkinson’s Disease without Dementia. IEEE/ACM Trans Comput Biol Bioinform. 2018;15(2): 570–580.

49. Kawashima S, Shimizu Y, Ueki Y, Matsukawa N. Impairment of the visuospatial working memory in the patients with Parkinson’s Disease: an fMRI study. BMC Neurol. 2021;21(1): 335.

50. Grossman M, Crino P, Reivich M, Stern MB, Hurtig HI. Attention and sentence processing deficits in Parkinson’s disease: the role of anterior cingulate cortex. Cereb Cortex. 1992;2(6): 513–525.

51. Suo X, Lei D, Li N, Peng J, Chen C, Li W, et al. Brain functional network abnormalities in parkinson’s disease with mild cognitive impairment. Cereb Cortex. 2022;32(21): 4857–4868.

52. Pereira JB, Aarsland D, Ginestet CE, Lebedev AV, Wahlund LO, Simmons A, et al. Aberrant cerebral network topology and mild cognitive impairment in early Parkinson’s disease. Hum Brain Mapp. 2015;36(8): 2980–2995.

53. Caspell-Garcia C, Simuni T, Tosun-Turgut D, Wu IW, Zhang Y, Nalls M, et al. Multiple modality biomarker prediction of cognitive impairment in prospectively followed de novo Parkinson disease. PLoS One. 2017;12(5): e0175674.

54. Liu G, Locascio JJ, Corvol JC, Boot B, Liao Z, Page K, et al. Prediction of cognition in Parkinson’s disease with a clinical-genetic score: a longitudinal analysis of nine cohorts. Lancet Neurol. 2017;16(8): 620–629.

55. Schrag A, Siddiqui UF, Anastasiou Z, Weintraub D, Schott JM. Clinical variables and biomarkers in prediction of cognitive impairment in patients with newly diagnosed Parkinson’s disease: a cohort study. Lancet Neurol. 2017;16(1): 66–75.

56. Kalaitzakis ME, Pearce RK. The morbid anatomy of dementia in Parkinson’s disease. Acta Neuropathol. 2009;118(5): 587–598.

57. Legault-Denis C, Aghourian M, Soucy JP, Rosa-Neto P, Dagher A, Aumont E, et al. Normal cognition in Parkinson’s disease may involve hippocampal cholinergic compensation: An exploratory PET imaging study with [(18)F]-FEOBV. Parkinsonism Relat Disord. 2021;91: 162–166.

58. Appel-Cresswell S, de la Fuente-Fernandez R, Galley S, McKeown MJ. Imaging of compensatory mechanisms in Parkinson’s disease. Curr Opin Neurol. 2010;23(4): 407–412.

59. Biundo R, Weis L, Antonini A. Cognitive decline in Parkinson’s disease: the complex picture. NPJ Parkinsons Dis. 2016;2: 16018.

60. Laansma MA, Bright JK, Al-Bachari S, Anderson TJ, Ard T, Assogna F, et al. International multicenter analysis of brain structure across clinical stages of Parkinson’s disease. Mov Disord. 2021.

